# Improving operative outcomes in patients with stomas

**DOI:** 10.64898/2026.01.05.26343439

**Authors:** Stella M. Dilke, Alistair Noble, Lydia R. Durant, Cristina Balcells, Jesus Miguens-Bianco, Julie A. K. McDonald, Nathan Danckart, Panagiotis Vorkas, Alexandros Siskos, James Willsmore, Anne L. McCartney, Hector C. Keun, George B. Hanna, Stella C. Knight, Philip J. Tozer, Ana Wilson, Carolynne J. Vaizey, Lesley Hoyles

## Abstract

Stoma formation diverts the flow of faeces to an opening on the skin, away from a section of bowel. Stomas are used to reduce the risks of bowel surgery and their effects are thought to be temporary and benign. However, negative effects [e.g. diversion colitis (inflammation in the excluded segment), poor quality of life, and an unclear effect on homeostasis] occur. We investigated these changes and whether distal feeding (DF) – the introduction of nutrition directly into the defunctioned bowel prior to stoma reversal – could mitigate these effects.

A series of clinical, qualitative, immunological, metabolomic and microbiomic experiments conducted across 133 patients identified consistent changes in immune and microbiome response which differed according to stoma formation and underlying disease process [colorectal cancer (CRC) with or without chemotherapy, intestinal failure (IF)], including a unique immune signature in circulating memory T-cell homing in stoma patients. DF induced the proliferation and altered homing of memory T cells, particularly in patients who had received chemotherapy. DF led to changes in serum metabolites, increased circulating markers of gut health (citrulline, serotonin), and modified the faecal microbiota. DF also promoted a faster return of bowel function and earlier hospital discharge in patients with CRC, while in IF it accelerated intestinal autonomy from parenteral nutrition.

This in-depth analysis of sequential experiments characterises gut homeostasis and post-operative systemic immunity after stoma formation in CRC and IF patients, and quantifies and explains the benefit of DF in these groups.

This research program has wide-ranging implications for patients worldwide with stomas. Given its simplicity, affordability and wide applicability, DF has the potential to become a transformative adjunct in surgical recovery and improved long-term gut health in patients with stomas.

## INTRODUCTION

Formation of stoma (FOS) is a common procedure in bowel surgery and is undertaken in many surgical circumstances, to reduce the risks of anastomotic leak in downstream bowel following anterior resection of colorectal cancer (CRC), to reduce ongoing inflammation (in inflammatory bowel disease) or as lifesaving damage control following a traumatic injury to the bowel.

Despite its ubiquity, FOS has been associated with increased short-term morbidity (1) including chronic dehydration (2) and acute kidney injury (3), and longer-term issues including inflammation in the excluded bowel (diversion colitis) (4). FOS has also been implicated as a risk factor for long-term pelvic floor dysfunction (Low Anterior Resection syndrome, LARS (5)) following reversal of stoma (ROS) in patients with CRC. Despite ROS frequently being a short and uncomplicated surgical procedure, patients can stay in hospital for up to 10 days post-operatively (6) with increased rates of wound infections and intolerance to oral intake of food (i.e. ileus). Post-operative ileus affects a quarter of patients undergoing bowel surgery for any reason, but the high prevalence following ROS is of particular interest to the surgical community (7), as prevention of ileus and management of other complications might facilitate earlier discharge from hospital, fewer complications and better quality of life for patients worldwide.

The effect of FOS on gut homeostasis and systemic immunity is poorly understood. The association between these effects and clinical outcomes, and strategies to reverse them and improve outcomes are the basis of this study’s series of experiments.

Distal feeding (DF) – the introduction of nutrition into a defunctioned bowel segment – has been demonstrated in studies in intestinal failure (IF) (8) to reduce the requirement for parenteral nutrition (PN) and increase small bowel mass. However, its broader application in CRC remains underexplored and the mechanism of action underlying its potential benefits is poorly understood (9).

In this study, we aimed to evaluate DF as a practical, patient-led intervention that could improve clinical outcomes such as restoration of bowel function and reduced length of hospital stay, assess its impact directly on patient quality of life and uncover the mechanistic impact of DF on systemic immunity, metabolites and the gut microbiota.

We performed a series of mechanistic, qualitative and clinical studies to identify the impact of both a stoma and DF in patients following CRC and IF surgery, alongside matched controls (n=133; including retrospective and prospective) to determine the plural impact of DF (**Figure 1A**). We used two DF diets: Vital (VITAL 1.5 CAL, a commercially available elemental feed) and chyme (recycled autologous enteric content).

**Figure 1.**
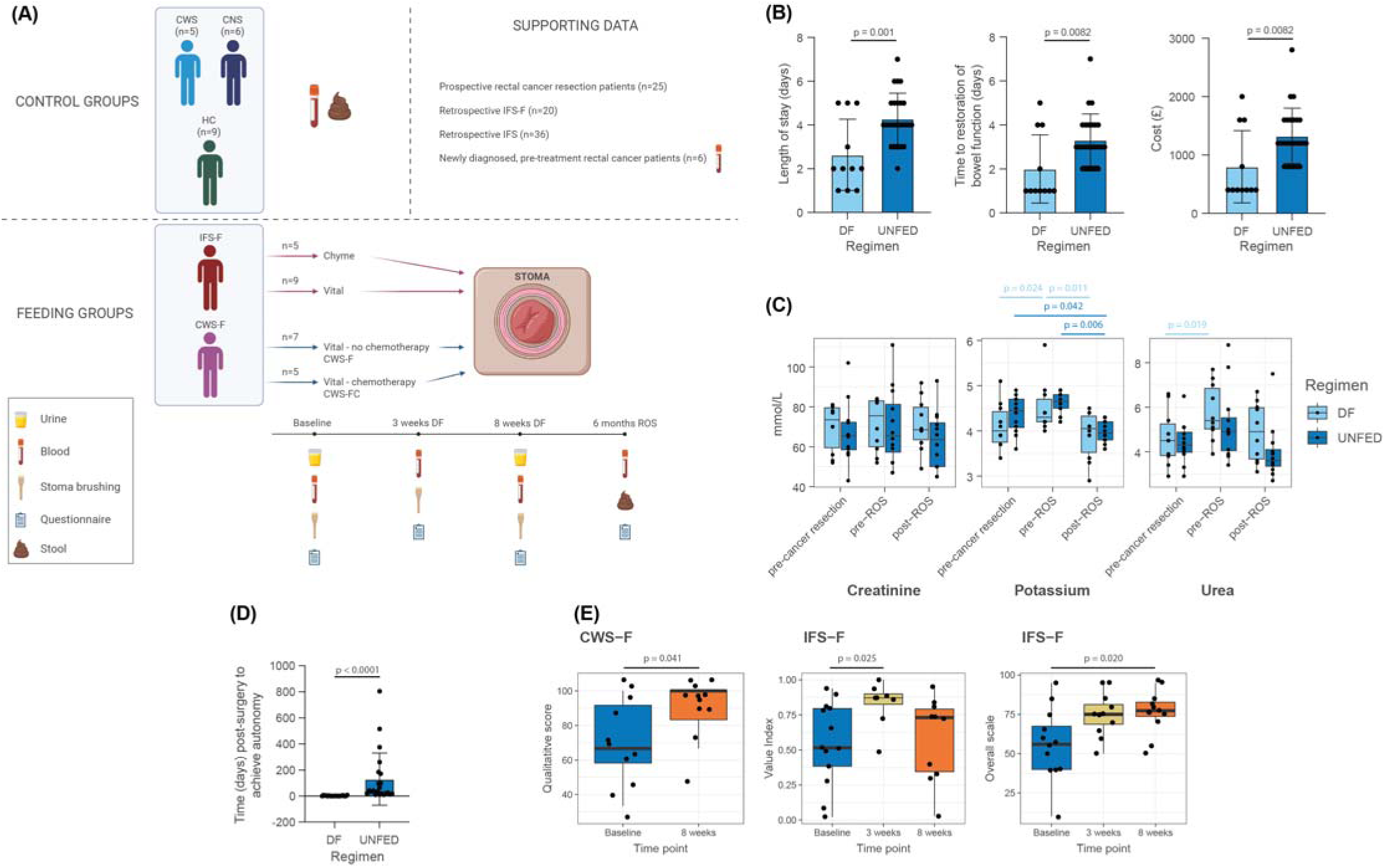
Study design and improvements in patient outcomes. (A) Study design showing the distal feeding (DF) and control groups used in this study. CWS, cancer with stoma; CNS, cancer no stoma; HC, healthy control; IFS-F, intestinal failure with stoma and DF; CWS-F, cancer with stoma and DF (no chemotherapy); CWS-FC, cancer with stoma, chemotherapy and DF. (B) DF significantly reduced length of time CRC patients stayed in hospital and led to significant reductions in healthcare-associated costs. DF, n=12 distally fed CRC patients; UNFED, n=25 CRC patients without DF). (C) Changes in renal function pre-cancer resection, prior to and post ROS in DF and non-DF patients. DF, n= 20; UNFED, n=35. D) DF significantly reduced time to intestinal autonomy in patients in IF patients. DF, n=10; UNFED, n=12. E) DF led to improvements in quality-of-life assessments for patients. CWS-F: Improvement in role performance in patients with CRC and DF across the 8-week regimen. IFS-F, left-hand panel: Improvement in qualitative role index in patients with DF and IF with greatest improvement at week 3 of the regimen. IFS-F, right-hand panel: Improvement in self-reported quality-of-life assessment across 8-week regimen in patients with IF and DF. (B-E) P values calculated using non-paired t tests (after normality testing).

## RESULTS

A central objective of this study was to assess whether DF provides clinically meaningful and patient-acceptable benefits. Without demonstrable advantages in real-world outcomes or tolerability, the value of DF as a therapeutic intervention would be limited, regardless of its mechanistic effects.

To evaluate the clinical benefit of DF in the CRC population, a prospective comparison was performed between patients who received DF prior to ROS (n=11) and contemporaneous controls who did not (n=25). All patients underwent a rectal cancer resection (total mesorectal excision (TME)) and ROS during the same year and at the same hospital site (**Figure 1A**). Clinical endpoints included length of inpatient stay and time to restoration of bowel function (defined as passage of flatus or stool). Patients with CRC in the DF group demonstrated a significantly shorter time to return of bowel function (mean reduction of 1 day; *p* = 0.008) and a reduced length of hospital stay (mean reduction of 1 day; *p* = 0.001) compared with non-DF controls (**Figure 1B**). This translated into a notable healthcare cost saving, estimated at £367 per inpatient day based on NHS coding data (**Figure 1B**).

To explore biochemical changes during the DF regimen, a sub-cohort of DF patients (n=10) underwent serum urea and electrolyte testing prior to rectal cancer resection, at 8 weeks of DF and again following ROS. These results were compared to non-DF CRC ROS controls (n=12) to assess the impact of ileostomy alone (**Figure 1C**).

DF patients showed an increase in serum potassium within normal range during the DF period (*p* = 0.02), which then decreased post-ROS (*p* = 0.01). In contrast, the non-DF group exhibited no significant change during ileostomy but showed a marked drop in potassium levels following ROS (*p* = 0.0006). This suggests a protective physiological effect of DF on electrolyte balance, with potential implications for mitigating acute kidney injury risk in this patient group. Serum urea increased during DF (*p* = 0.02), without associated changes in serum creatinine, indicating a metabolic response to enteral stimulation.

In patients with IF, the timing of ROS and heterogeneity of underlying pathology limited the feasibility of prospective comparison (other than longitudinally). A retrospective cohort study was therefore conducted. Twenty patients who received DF prior to ROS were identified and compared to a matched control group of 35 patients who did not receive DF. Groups were balanced with respect to age, sex, aetiology of IF, duration of PN, and pre-operative small bowel length (**Supplementary Table 1**).

DF significantly increased the likelihood of achieving intestinal autonomy, defined as complete independence from PN (11) (DF group: 95%, 19/20 vs control: 60%, 21/35; *p* < 0.01). Furthermore, time to autonomy was significantly shorter in the DF group (*p* = 0.03) (**Figure 1D**), suggesting that DF facilitates recovery of intestinal absorptive function.

Patient experience and acceptability were evaluated using both qualitative interviews and validated quality-of-life instruments. Eighteen semi-structured interviews were conducted with CRC (n=11) and IF (n=7) patients during their DF regimen. Thematic analysis revealed key recurring themes: patients felt empowered by DF, reporting improvements in energy, wellbeing, weight gain, and a sense of agency in their recovery. The importance of a robust support network was highlighted as a key facilitator of adherence.

Quantitative quality-of-life was assessed using the EQ-5D-5L for all patients at all timepoints, and EORTC-QLQ-C30 and QLQ-C29 for CRC patients, including after ROS. CRC participants demonstrated significant improvements in role functioning during the DF period (difference in means = 19.70 ± 9.02, 95% CI 0.89 to 38.50; *p* = 0.04), suggesting enhanced capacity to engage in daily activities. In the IF group, significant improvements in overall quality of life were observed by week 3 of DF across both EQ-5D metrics: patient self-rating (*p* = 0.02) and index-based score (*p* = 0.025) (**Figure 1E**).

### DF promotes proliferation and tissue-homing of circulating adaptive immune memory cells

To investigate the immunological mechanisms underlying the clinical effects of DF, we performed longitudinal analyses of circulating T and B cell responses in patients with CRC and IF. We hypothesised that DF would modulate systemic immune memory through exposure of the distal gut to luminal antigen, restoring immune homeostasis altered by stoma formation.

Peripheral blood mononuclear cells (PBMCs) were isolated from participants at four time points: baseline (pre-DF), 3 weeks of DF, 8 weeks of DF, and 6 months post-ROS (CRC group only) (**Figure 1A**). Cells were cultured and stimulated *in vitro* with a panel of 19 gut commensal bacterial antigens (**Supplementary Table 2**) (10). Proliferative responses of CD3^+^/CD4^+^ T cells, CD3^+^/CD8^+^ T cells, and CD19^+^ B cells were assessed using cell trace violet staining and flow cytometry. These data were compared longitudinally throughout the DF regimen, and directly with healthy and unhealthy controls.

In patients with CRC, DF significantly enhanced *in vitro* proliferative responses in all adaptive immune cell subsets assessed. After adjusting for background proliferation, the predominant response was observed in CD3^+^ /CD4^+^ T cells, showing a significant increase between 3 and 8 weeks of DF (*p* = 0.01) and from baseline to 8 weeks (*p* = 0.006), indicating a time-dependent immunostimulatory effect (**Figure 2A**).

**Figure 2.**
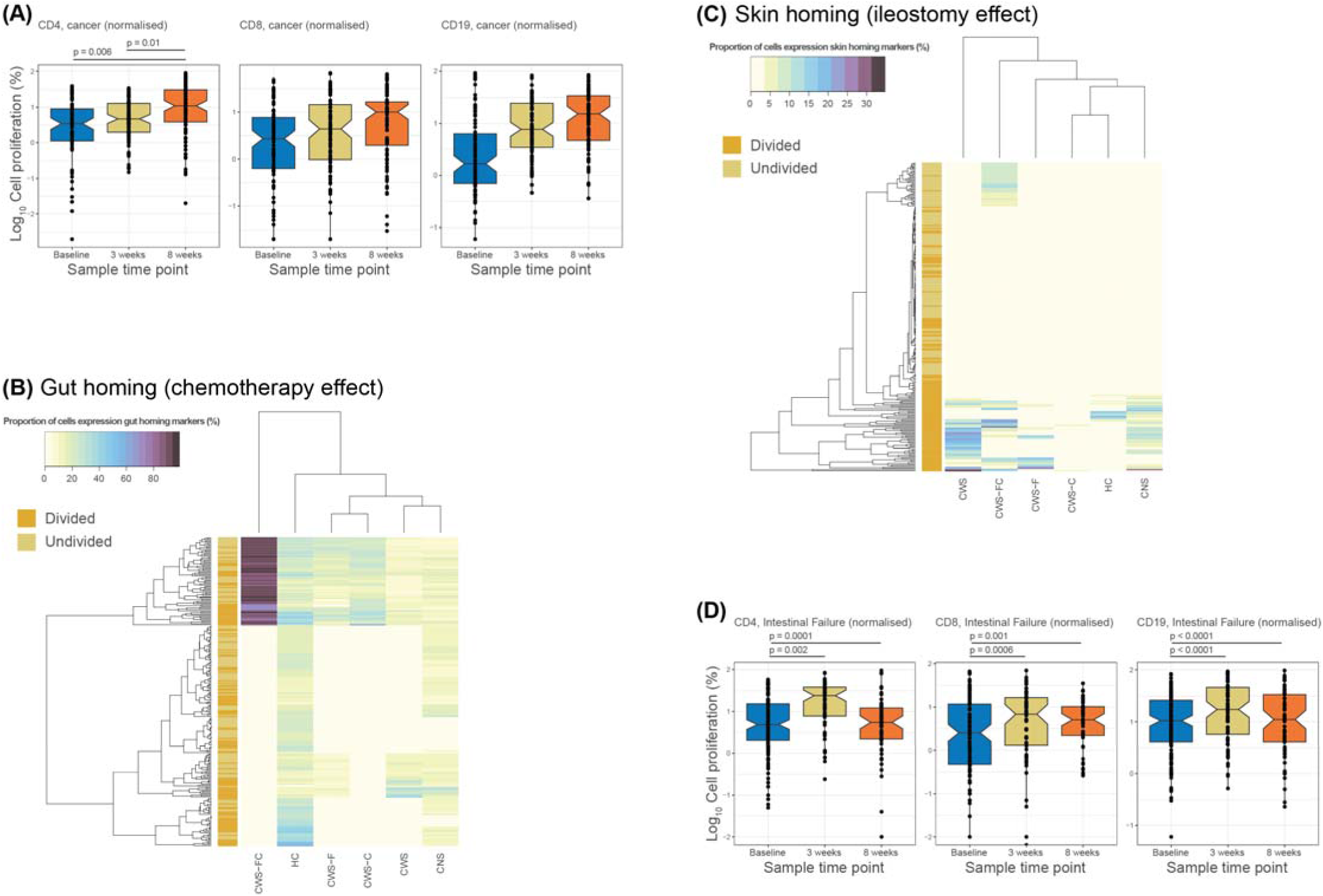
Circulating PBMC responses to DF. (A) DF has a mechanism grounded in circulating adaptive immune memory. Normalised cell proliferation rates of *in vitro* CD4^+^/CD3^+^ cells over the 8-week DF regimen in patients with treated CRC, with increased proliferation after 8 weeks of DF. (B) Heat map demonstrating gut homing within the CRC ROS DF groups, compared to healthy and non-healthy controls: CWS–FC, cancer with stoma, DF and chemotherapy (n=2); HC, healthy control (n=7); CWS–F, cancer with stoma, DF (n = 3); CWS-C, cancer with stoma, chemotherapy (n=3); CWS, cancer with stoma (n=2); CNS, cancer no stoma (n=6). Top left-hand corner shows gut homing in CWS–FC patients. (C) Heat map demonstrating skin homing within CRC ROS DF groups, compared to healthy and non-healthy controls: CWS–FC, n= 2; HC, n=7; CWS–F, n=3; CWS-C, n=3); CWS, n= 2; CNS, n=6. Heat map shows increased skin homing in patients with stomas, particularly in divided cells. (B, C) P values were calculated with one way ANOVA. (D) The right-hand panel demonstrates increased proliferation in patients with IF in all cell groups tested after 3 weeks of the DF regimen. Data are longitudinal and were analysed with Friedman ranking due to non-parametric distribution.

These responses were phylum-specific. CD4^+^ cell proliferation was significantly increased in response to *Actinomycetota* (*p* = 0.01 at 8 weeks), *Pseudomonadota* (*p* = 0.01), and *Bacillota* (*p* = 0.03), suggesting broad reactivation of gut-specific memory across diverse bacterial taxa. Notably, no increase in CD4^+^ responses to *Bacteroidota* was observed.

CD8^+^ and CD19^+^ cell proliferation was also increased, but more limited in scope. Both subsets showed enhanced responses to *Pseudomonadota* (*p* = 0.01 for CD8^+^ at 3 weeks and CD19^+^ at 8 weeks). CD19^+^ cells demonstrated progressive reactivity to *Staphylococcus epidermidis* over the course of DF (*p* = 0.04 at 8 weeks) (**Supplementary Figure 1**).

Stratified analysis revealed significantly abnormal circulating immune responses in CRC patients who had received chemotherapy. CD4^+^ T cells from chemotherapy-treated patients demonstrated an exaggerated response to *Bacteroides* ovatus between baseline and 3 weeks (*p* = 0.02), while CD8^+^ T cells exhibited reduced responses to *Veillonella atypica* between baseline and 8 weeks (*p* = 0.03) (**Supplementary Figure 2**).

Homing marker expression was also measured on circulating T and B cells by flow cytometry examining both proliferated (divided, *in vitro*) and non-proliferated (undivided, *in vivo*) PBMCs in patients post-DF and ROS.

Both CD4^+^ and CD8^+^ T cells from CRC patients with ROS, chemotherapy and DF displayed high expression of the gut-homing marker integrin β7 in response to bacterial stimulation at 6 months following ROS (*p* = 0.0001). This exaggerated gut-homing marker expression was not observed on T/B cells from patients with CRC and ROS, patients with CRC, ROS and chemotherapy alone, CRC without stoma or in healthy controls, suggesting that DF partially reprogrammes tissue-specific immune trafficking of circulating lymphocytes in the context of prior cytotoxic therapy, directing cells towards to the gut (**Figure 2B**).

T/B cells from patients who underwent DF prior to reversal in both IF and CRC (without chemotherapy) showed a similar incremental increase in proportion of cells expressing integrin β7 in response to bacterial stimulation throughout the 8-week regimen. In CRC (assessed additionally after ROS) this returned to baseline, suggesting DF may play a role in resetting aberrant tissue-specific immune targeting induced by stoma formation.

Interestingly, DF patients exhibited a similar proportion of T/B cells expressing the skin homing marker CLA in both divided and undivided cells, whereas post-ROS CRC patients (without prior DF) there was a higher proportion of CLA-expressing T/B cells in divided cells (i.e. *in vitro*) only (**Figure 2C**), suggesting that DF dampens the longer-term impact of a stoma by redirecting immune cells away from the skin/to the gut.

In patients with IF, DF induced widespread systemic immune activation. Unadjusted analyses of blood PMBCs showed significant increases in CD4^+^, CD8^+^, and CD19^+^ proliferation by 3 weeks of feeding (*p* < 0.0001). After background correction, peak antigen-specific proliferation occurred at 3 weeks across all three cell types, suggesting a temporally concentrated immune response to luminal re-exposure, occurring at an earlier timepoint than in CRC (**Figure 2D**).

Unlike the CRC group, antigen specificity in IF patients was less clearly delineated, with phylum-level significance only noted in *Bacillota* (*p* = 0.01) (**Supplementary Figure 3**). These findings suggest that DF in IF patients induces a generalised immune activation, potentially reflecting prolonged immune dormancy or mucosal immune dysregulation secondary to long-term stoma use (i.e. diversion colitis).

### DF elicits long-term changes to the faecal but not the mucosal microbiota

It is important to reiterate here that the CRC patients were free of cancer at the time of microbiota sampling (at ROS and 6 months after ROS). It was expected that DF would drive changes in gut microbial communities: mucosal samples were taken longitudinally using ileal brushes; faecal samples were collected 6 months after ROS. Results from the metataxonomic (i.e. 16S rRNA gene-based) analysis of ileum mucosal samples showed no differences in microbial abundances or alpha/beta diversity metrics across DF timepoints in this area of the gut nor at ROS (data not shown).

Shotgun metagenomic data were generated from faecal samples taken 6 months after ROS for distally fed CRC patients, and control samples with or without ileostomy were matched according to their timeframe (same calendar year, similar approximate time of ileostomy closure and matched for age and chemotherapy status) (**Figure 3**).

**Figure 3.**
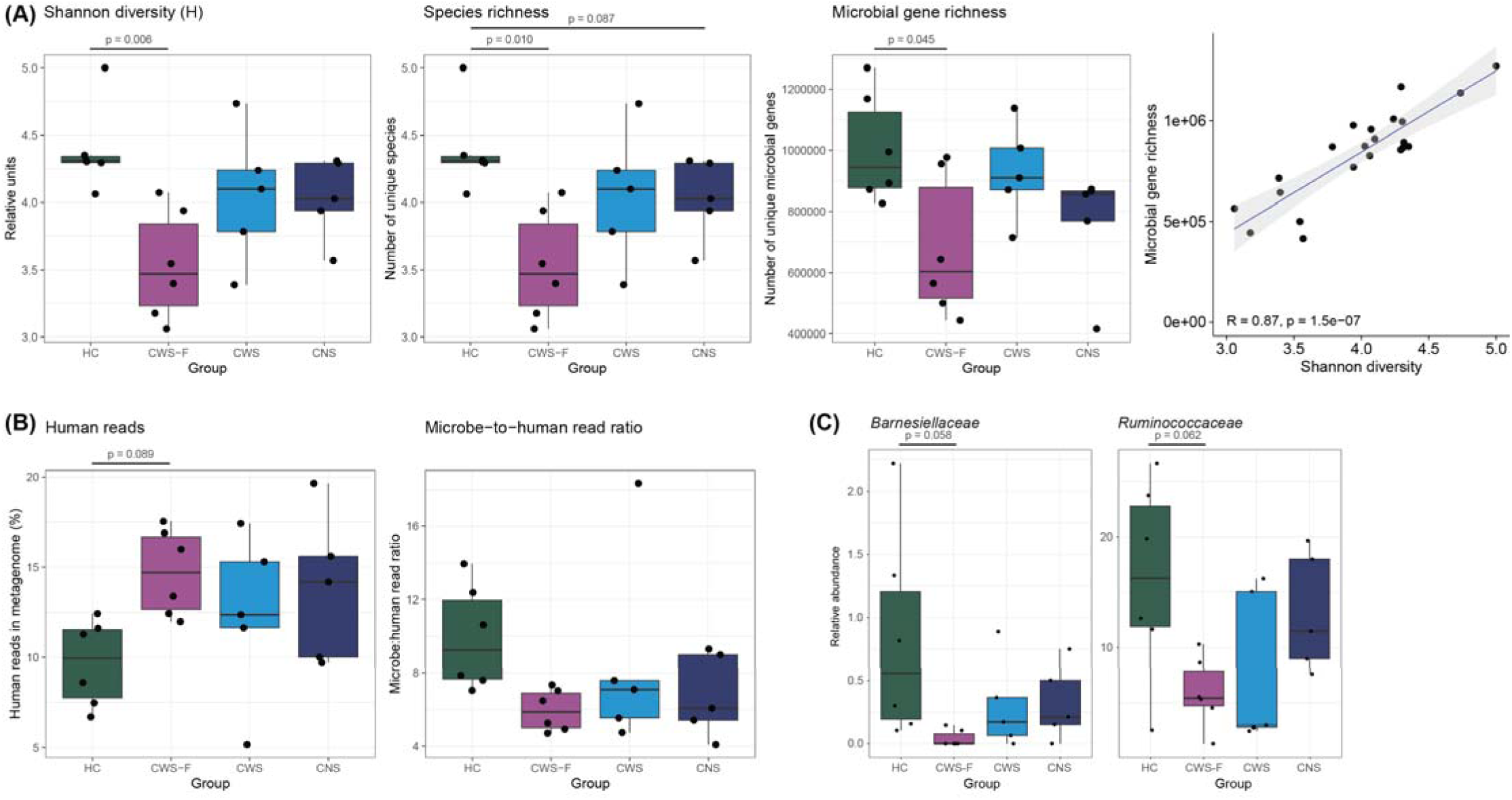
Assessment of the effects of DF and/or stoma on the faecal microbiota of patients 6 months after ROS. (A) Assessment of various alpha diversity metrics demonstrated the faecal microbiota of cancer patients was different from that of healthy controls. (B) Consideration of human reads in metagenomic data and inferences based on non-human-to-human read ratios (non-human read count divided by human read count) predicted cancer patients harboured lower bacterial biomass in their faecal microbiota compared with healthy controls. (A–C) HC, healthy control (n=6); CWS-F, cancer with stoma and DF (n=6); CWS, cancer with stoma (no DF) (n=5); CNS, cancer no stoma (n=5). Two-way ANOVA with Tukey HSD testing (multiple pairwise-comparison between the means of groups) was used to determine statistically significant differences when considering feeding regimen, group affiliation or stoma status (significant adjusted p values given in main text). One-way ANOVA with Tukey’s post hoc testing was used to compare groups to one another (significant adjusted p values shown on images).

The faecal microbiota of metabolically healthy adults has higher microbial species and gene richness than adults affected by inflammation-associated conditions such as obesity, insulin resistance, dyslipidaemia and steatotic liver disease (12)(13)(14). Comparison of species-and gene-richness metrics for the faecal microbiota of individuals included in this study demonstrated healthy controls had significantly higher alpha diversity scores (Shannon diversity FDR-*p*_adj_ = 0.010; species richness FDR-*p*_adj_ = 0.005) and microbial gene richness (FDR-*p*_adj_ = 0.028) when compared against all cancer patients, irrespective of feeding group or whether the cancer patients previously had a stoma (**Figure 3A**). It was notable that the DF cancer patients’ faecal microbiota never returned to a microbiota comparable to that of healthy individuals with respect to species and gene richness.

Consideration of the ratio of bacterial to human reads contributing to metagenomes has recently been proposed as a means of normalizing metagenomic data to predict bacterial biomass, with ratios varying across host health and disease states (15). Former cancer patients (all groups) had significantly (FDR-*p*_adj_ = 0.025) higher levels of human DNA in their metagenomes than healthy controls (**Figure 3B**). Our previous work has shown limited contribution of non-bacterial reads to faecal metagenomic data (14). As such, we consider the ratio of non-human reads to human reads to be comparable to the bacterial-to-human-read ratio. Across all cancer groups the ratio of bacteria to human reads was significantly reduced (FDR-*p*_adj_ = 0.083) compared with healthy individuals (**Figure 3B**), suggesting that patients now free of cancer have greatly reduced gut bacterial biomass compared with healthy individuals who have never had cancer.

No significant taxon-specific changes in abundance data were noted from phylum to order level for the metagenomic data. Significant (FDR-*p*_adj_ <0.1) family-level differences for two taxa of anaerobic bacteria were observed: *Barnesiellaceae* and *Ruminococcaceae* were present in lower abundance in cancer patients than healthy controls (**Figure 3C**). The genus Barnesiella was also significantly (FDR-*p*_adj_ = 0.058) less abundant in the DF-fed stoma patients compared with healthy controls (**Supplementary Figure 4**).

Sample sizes did not afford comparisons of DF cancer patients with stoma and whether they had had chemotherapy. However, given our findings in relation to immune responses, chemotherapy, anaerobe abundance and bacterial biomass in former cancer patients compared to healthy individuals, dual and full consideration of changes in immune responses (and reactive oxygen species) and gut microbiota dynamics are required in future mechanistic studies focused on post-cancer recovery and/or DF.

### DF influences circulating serum metabolites and validates chyme as a superior DF regimen

To understand the systemic impact of DF on circulating metabolites, longitudinal serum metabolomic profiles were determined using liquid chromatography mass spectrometry (LC-MS) in nine CRC patients (four of whom had undergone chemotherapy). Additionally, targeted analyses were performed in the IF cohort comparing patients fed with Vital versus chyme. The aim was to determine whether DF altered key metabolic pathways, and whether the feed composition influenced these (**Figure 4**).

**Figure 4.**
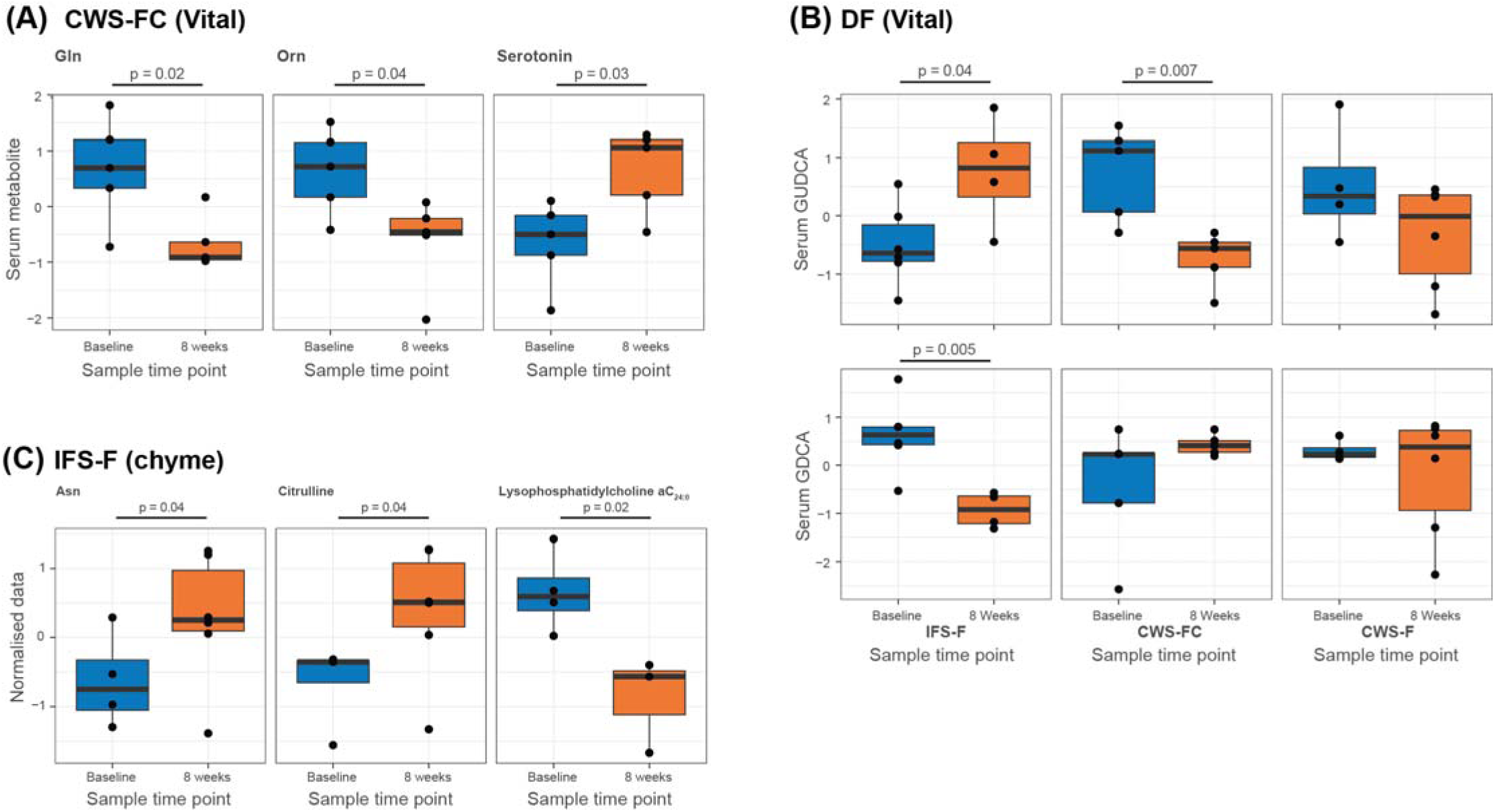
Effects of DF on the serum metabolome of CRC and IF patients. (A) Demonstrates the impact of Vital 1.5 in patients with CRC, DF and chemotherapy. Serum plasma showed a reduction in glutamine and ornithine and an increase in serotonin over the 8-week regimen (n=5). This is not seen in the CRC, DF and non-chemotherapy group, nor in the DF IF patient group used as controls. (B) Demonstrates the impact of Vital 1.5 in patients with IF on secondary bile acids (GUDCA, GDCA). (C) Demonstrates the impact of chyme feeding in patients with IF. Increases were seen in citrulline, a marker of small bowel mass, and demonstrates the superiority of chyme as a DF diet when compared with Vital 1.5.

In CRC patients who had received chemotherapy, DF significantly reduced circulating levels of glutamine and ornithine from baseline to 8 weeks (**Figure 4A**). These amino acids are essential for enterocyte function and immune cell proliferation (16), and their depletion may reflect increased metabolic utilisation or shifts in gut microbial metabolism.

Chemotherapy patients also exhibited significant increases in serotonin during DF, a key neuromodulator produced both by enterochromaffin cells and certain anaerobic microbes (17). Elevated serotonin has been linked to improved gut barrier integrity and neuronal signalling (18), supporting a possible role for DF in promoting gut health in this group of patients (19).

Of note, chemotherapy patients had reduced levels of serum glycoursodeoxycholic acid (GUDCA) during DF (**Figure 4B**), a secondary bile acid with known anti-inflammatory properties (20). This reduction was not observed in non-chemotherapy or IF patients, suggesting impaired microbial bile acid metabolism in the chemotherapy group.

In the IF cohort, choice of DF diet had a significant impact on metabolic outcomes. Patients receiving chyme DF demonstrated a significant increase in serum citrulline levels over the 8-week regimen (*p* = 0.04) (**Figure 4C**). As citrulline is a validated biomarker of functional small bowel mass (21) this finding supports chyme DF as a method to promote intestinal regeneration in patients with IF.

### DF reduced inflammation in defunctioned bowel on histopathological analysis

Eighteen patients had pre-feeding histopathology samples taken via endoscopy. Of these, two CRC patients had evidence of diversion colitis on histopathology (**Figure 5**). Two patients had evidence of atrophy (1 CRC, 1 IF). At 8 weeks of DF, both instances of diversion colitis, and one patient with atrophy had complete histological resolution and were independently reported as normal ileal biopsies.

**Figure 5.**
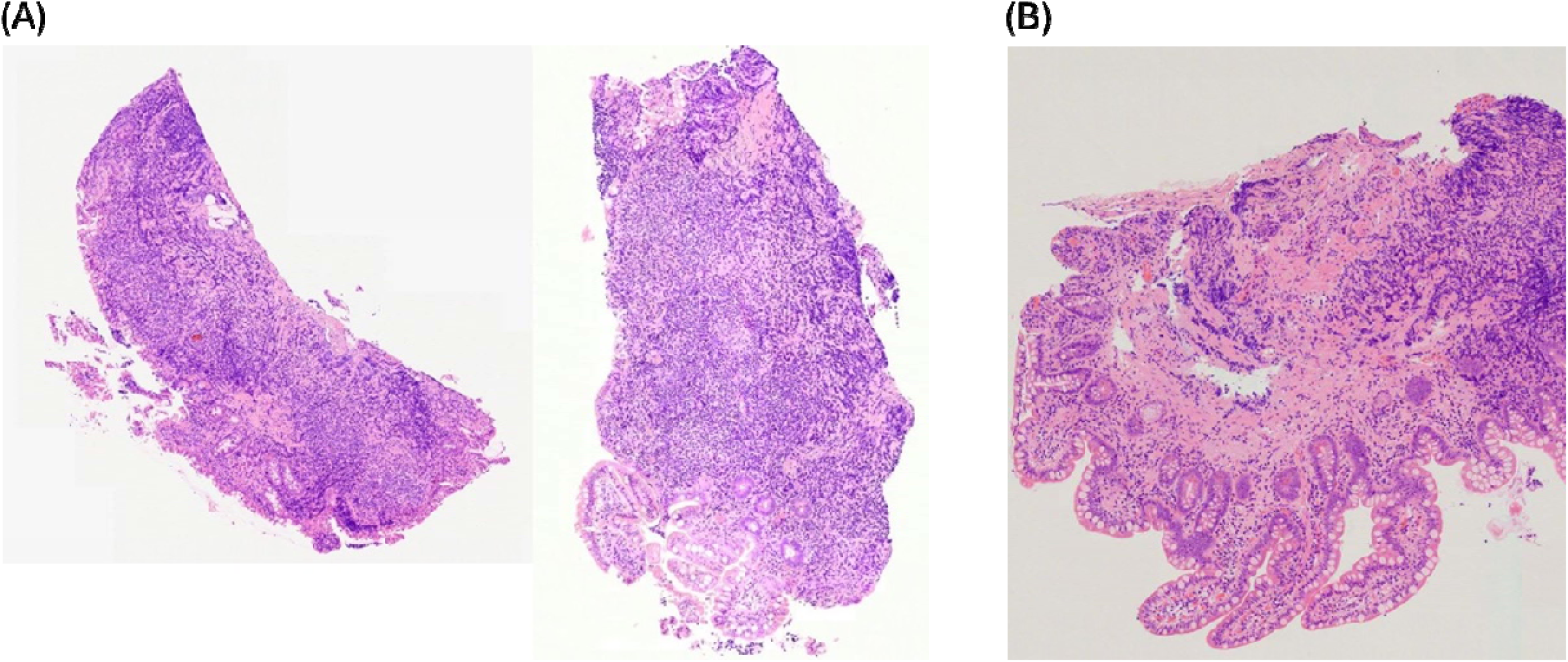
Effect of DF on diversion colitis in a colorectal cancer patient. (A) Pre-DF the patient had evidence of atrophy and diversion colitis. Note the flattened villi in both images and inflammatory deposits in the centre of both images. (B) At 8 weeks DF, the same patient had a complete resolution of diversion colitis. Villi were evident and inflammatory deposits were absent.

## DISCUSSION

This study challenges the long-held assumption that defunctioning stomas are physiologically benign. Through a series of clinical, qualitative, immunological, metabolomic, and microbiome analyses, we demonstrate that patients with stomas – whether for CRC or IF – exhibit persistent systemic immune abnormalities and altered gut physiology. These changes persist even after restoration of bowel continuity and are particularly pronounced in patients who have undergone chemotherapy. Our data show that DF is a simple, safe, patient-led intervention that improves perioperative outcomes, reduces healthcare-associated costs, modulates systemic immunity, and exerts measurable metabolic and microbiome effects.

Clinically, DF was associated with significantly shorter hospital stays and faster return of bowel function following reversal of ileostomy in CRC patients. In IF patients, DF increased the likelihood of achieving intestinal autonomy and reduced the time to PN independence. These outcomes not only reflect improved bowel function but also suggest meaningful cost savings and reductions in post-operative morbidity. Qualitative analysis confirmed that patients perceived DF as a proactive and empowering intervention that improved energy levels, appetite, and overall well-being. Together, these data support DF as a viable pre-operative strategy to improve surgical readiness and recovery.

In cancer patients, changes related to the disease itself or chemotherapy, rather than FOS, were not reversible. Our microbiome analyses show that the gut microbial ecosystem in CRC patients remains profoundly altered even after cancer cure and restoration of bowel continuity. Faecal microbiota in DF patients did not regain the richness or functional gene diversity observed in healthy controls, and biomass estimates (via bacterial:human DNA ratios) suggest a substantially reduced microbial load in cancer patients. These changes were independent of whether patients had a stoma, indicating that CRC – and perhaps its treatments – leave a lasting ecological imprint on the gut microbiota. Specific reductions in families such as *Ruminococcaceae* and *Barnesiellaceae* further support a loss of obligate anaerobes, which may have implications for immune modulation, barrier integrity, and long-term colorectal health.

Chemotherapy-exposed patients were uniquely abnormal in their gut homing responses, with significantly heightened expression of integrin β7 on T cells up to six months after stoma reversal. These findings suggest a long-lasting imprint of chemotherapy on mucosal immune regulation, which may contribute to chronic bowel dysfunction after cancer treatment. The immune proliferation responses observed during DF were broad, suggesting that DF acts as a general immunological stimulus, rather than promoting narrow, species-specific reactivity.

However, FOS induced a characteristic effect on gut homeostasis and systemic immunity that DF was able to mitigate. Both CRC and IF patients with stomas exhibit aberrant memory T and B cell responses to gut commensals, with altered antigen-specific proliferation and tissue-homing signatures. Importantly, DF drives a transient but robust proliferation of circulating memory cells, particularly CD4^+^ and CD19^+^ cells, and normalises key features of mucosal immune activation.

Metabolomic profiling revealed additional mechanistic insights. CRC patients receiving DF showed significant alterations in amino acid metabolism, including decreased serum glutamine and ornithine, particularly in those who had received chemotherapy. These findings may reflect both microbial and host utilisation of key metabolic intermediates. Chemotherapy patients also had elevated circulating serotonin, a marker associated with improved gut function and microbial activity (22).

Chyme-based DF, as used in the IF cohort, was particularly effective in increasing serum citrulline, a recognised marker of enterocyte mass and mucosal health (23). This reinforces the superiority of chyme over formula-based DF diets in promoting intestinal adaptation. However, self-administration of this diet was conceptually unacceptable to our CRC patients. Adoption of automated chyme reinfusion devices for use in these patients should be a focus of future studies.

Taken together, our findings redefine the physiological impact of temporary stomas and highlight a previously underappreciated window for intervention. The abnormal immune, microbial and metabolic profiles identified here suggest that the period of stoma formation is not inert but pathophysiologically dynamic. DF represents a targeted opportunity to modulate gut–immune crosstalk, improve perioperative outcomes, and potentially alter long-term sequelae of bowel surgery and chemotherapy. Given its safety, simplicity, and patient acceptability, DF warrants further investigation as a scalable adjunct in surgical care, especially in the post-cancer setting. Future studies should focus on optimising DF dietary regimens, exploring its effect on innate immunity at the mucosal barrier, and determining its long-term impact on cancer recurrence, bowel function, and quality of life.

## FUNDING

Experiments performed by SMD were funded by Bowel Research UK (previously Bowel & Cancer Research) for the entirety of this work as part of their small grant funding, under the reference ‘*The impact of enteric feeding of the excluded terminal ileum and/or colon – mechanistic, qualitative and clinical aspects*’. LH was funded by UK MED-BIO, which was supported by the Medical Research Council (grant number MR/L01632X/1). LH and HCV were funded by the European Union’s Horizon 2020 research and innovation programme under grant agreement no. 874583. This publication reflects only the authors’ view and the European Commission is not responsible for any use that may be made of the information it contains. Funding for AN was from London North West University Healthcare NHS Trust R&D.

## Supporting information

Supplementary Material (including Methods)

## Data Availability

Shotgun metagenomic sequence data reported on this article are available from BioProject PRJEB38258. The metataxonomic (16S rRNA gene) data described in this study are available under BioProject PRJEB89732.

https://figshare.com/projects/_b_Improving_operative_outcomes_in_patients_with_stomas_b_/121884

## ACKNOWLEDGEMENTS

We would like to thank Suzanne Donnelly, Simon Gabe, Sarita Dhamala-Bhatta and Joy Odita for their input in patient recruitment, and in implementation of the DF pathway at St Mark’s Hospital. Thanks to Mia Small who aided sample collection. Adam Stearns at Norfolk and Norwich Hospital assisted with early idea synthesis and with inspiration for the qualitative work. Thanks to Dr Morgan Moorghen who analysed the histopathology samples. Bowel Research UK funded SMD’s work in full and were extremely helpful throughout the study, including during the COVID-19 pandemic. The study was also included in the NIHR portfolio which enabled research nurse input for control recruitment, which we are extremely grateful for. We would like to thank the St Mark’s Academic Institute for their input throughout the study. Additional thanks to Julie Hoong, Phebe Ekrebesi, Toby Pring, Sophie Zemenides and Alison Scoggins at the Antigen Presentation Research Group for their help enabling experiments throughout the course of the study, and their input at weekly research meetings. Mike Cox and Leah Cuthbertson are thanked for providing guidance on the processing of low-biomass microbiota samples in the metataxonomic analysis.

## AUTHOR CONTRIBUTIONS

SMD, CJV, AW, PJT, AN, SCK and LH designed the study. CJV is the clinical lead and LH is the scientific lead. PJT, AW, CJV, LH and GBH gave input on the direction of the work throughout, including input on ethics, patient recruitment and appropriate timescales for sample collection. SMD recruited all patients and collected all samples, other than the pre-resection samples which were collected by AN. JW and SMD analysed the retrospective IF data. SMD analysed all CRC retrospective data. CJV, AW and PJT advised weekly on patient groups and appropriate comparisons and liaised with other clinical teams. HCK supervised the metabolomic experiments and prepared samples for analysis with SMD. CBN and PV analysed the metabolomic data with input from AS and HCK. SMD extracted DNA from all samples with LRD and JMB. JMB and ND supervised the 16S rRNA gene sequencing and analysis, analysed the 16S rRNA gene sequence data and prepared samples with SMD, supervised by JAKM. ND advised on PCR and DNA preparation. LC advised on trace DNA analysis and made a reagent for extraction. LH processed and analysed the shotgun metagenomic data and interacted with all the other scientific teams within this study with SMD. LH and ALM provided microbial samples for analysis in the immune work. SMD cultured immune cells and analysed them supervised by LRD, AN and SCK. SCK gave weekly input on immunology results. SMD and LH wrote the manuscript with input from LRD, PJT, AW, AN and CJV. All authors approved the final version of the manuscript.

## COMPETING INTERESTS

None to declare.

## Abbreviations

CRC: colorectal cancer
DF: distal feeding
IF: intestinal failure
FOS: formation of stoma
ROS: reversal of stoma
PBMC: peripheral blood mononuclear cell
PN: parenteral nutrition.

## Notes

### Competing Interest Statement

The authors have declared no competing interest.

### Funding Statement

Bowel Research UK (previously Bowel & Cancer Research).
UK MED-BIO (grant number MR/L01632X/1).
European Union Horizon 2020 research and innovation programme under grant agreement no. 874583.
London North West University Healthcare NHS Trust R&D.

### Author Declarations

Research and Development at London North West University Trust and the UK National Research Ethic Centres (IRAS 248290) gave ethical approval for this work.

