## Supplementary Material (including Methods) for "Improving operative outcomes in patients with stomas"

**Supplementary Table 1.** Demographic comparisons between distally fed IF patients and matched controls

|  | **Distal feeding (DF)** | **No distal feeding (non-DF)** |
| --- | --- | --- |
| **Number** | n=20 | n=36 |
| **Sex** | M=11 F=9 | M=21 F=15 |
| **Sex ratio (M:F)** | 1.2 | 1.4 |
| **Age at continuity surgery (years)** |  |  |
| **Mean** | 50.7 | 54.0 |
| **Standard deviation** | 18.3 | 15.8 |
| **Range** | 16.3 to 77.8 | 21.9 to 85.7 |
| **IF aetiology (n, %)** |  |  |
|  | Crohn's 7 (35.0%) | Crohn's 6 (16.7%) |
|  | Surg complications 8 (40.0%) | Surg complications 14 (38.9%) |
|  | Mesenteric vascular disease 5(25.0%) | Mesenteric vascular disease 14 (38.9%) |
|  |  | Radiation enteritis 1 (2.8%) |
|  |  | Trauma 1 (2.8%) |
| **Proximal anatomy pre continuity surgery** |  |  |
|  | Jejunostomy 20 (100%) | Jejunostomy 36 (100%) |
| **Distal anatomy pre surgery** |  |  |
|  | Distal SB out of continuity 20 (100%) | Distal SB out of continuity 17 (47.2%) |
|  | Colon out of continuity 18 (90%) | Colon out of continuity 33 (91.7%) |
| **Pre-operative SB length from DJ flexure to stoma (cm)** |  |  |
| **Mean** | 50.1 | 74.5 |
| **Standard deviation** | 31.7 | 48.8 |
| **Range** | 0 to 225 | 0 to 165 |
| **Pre-operative SB length out of continuity (cm)** |  |  |
| **Mean** | 114.6 | 59.9 |
| **Standard deviation** | 76.1 | 49.5 |
| **Range** | 13.0 to 280.0 | 0 to 190.0 |
| **Duration HPN to continuity surgery (months)** |  |  |
| **Mean** | 9.5 | 7.1 |
| **Standard deviation** | 10.3 | 7.1 |
| **Range** | 2.5 to 42.4 | 0.7 to 37.7 |
| **Duration distal feeding prior to continuity surgery (months)** |  |  |
| **Mean** | 5.9 | Not applicable |
| **Standard deviation** | 3.7 | Not applicable |
| **Range** | 0.3 to 15.1 | Not applicable |
| **Volume distal feeding (ml/day)** |  |  |
| **Mean** | 183.3 | Not applicable |
| **Standard deviation** | 53.7 | Not applicable |
| **Range** | 100 to 300 | Not applicable |
| **Distal feed type** |  | Not applicable |
| **Distal feeding complications (n, %)** | 3 (13.6%) | Not applicable |
|  | 2 nausea, 1 nausea and vomiting | Not applicable |
| **Stopped distal feeding due to complications (n, %)** | 1 (4.5%) | Not applicable |
|  | 1 nausea | Not applicable |
| **Readmissions with complications of distal feeding** |  | Not applicable |
| **Length of stay following continuity surgery (days)** |  |  |
| **Mean** | 18.1 | 25.1 |
| **Standard deviation** | 13.7 | 17.4 |
| **Range** | 3 to 66 | 5 to 83 |
| **Anatomy post continuity surgery** |  |  |
|  | Jejuno-colic anastamosis 20 (90.9%) | Jejuno-colic anastamosis 33 (91.7%) |
|  | Jejunostomy/ileostomy 2 (9.1%) | Jejunostomy/ileostomy 1 (2.8%) |
|  |  | Ileo-anal pouch 2 (5.6%) |
| **Post-operative SB (cm)** |  |  |
| **Mean** | 186.5 | 100.1 |
| **Standard Deviation** | 96.9 | 69.2 |
| **Range** |  |  |
| **Achieved intestinal autonomy (n, %)** | 22 (95.7%) | 21 (58.3%) |
| **Anatomy post continuity surgery of those achieving autonomy** |  |  |
|  | Jejuno-colic anastamosis 20 (90.9%) | Jejuno-collic anastamosis 19 (90.5%) |
|  | Ileostomy 2 (9.1%) | Ileostomy 1 (4.8%) |
|  |  | Ileo-anal pouch 1 (4.8%) |
| **Post-operative SB (cm)** |  |  |
| **Mean** | 186.5 | 131.9 |
| **Standard deviation** | 96.9 | 64.0 |
| **Range** | 80.0 to 340.0 | 40.0 to 180.0 |
| **If achieved intestinal autonomy, time to achieve post continuity surgery (months)** |  |  |
| **Mean** | 2.4 | 3.3 |
| **Standard deviation** | 2.7 | 4.5 |
| **Range** | 0.0 to 8.1 | 10 to 375 |
| **Patients with post-operative complication/surgical post-operative complication** | 9 (40.9%)/6 (27.2%) | 16 (44.4%)/12 (33.3%) |
| **Readmission with surgical complication (time from surgery to readmission in months)** | 2 (10%) | 3 (8.3%) |
|  | Incisional hernia (30.3) | Abdo pain conservatively mx (3.2) |
|  | Abdo pain conservatively mx (11.5) | ECF repair (4.8) |
|  |  | SBO requriring relaparotomy (12.0) |
| **Mortality by end of follow up period** | 0 (0%) | 3 (8.%) |
|  |  | (at 16.6 months, 30.6 months, 6 months) |
| **Study follow up period (months)** |  |  |
| **Mean** | 75.3 | 58.9 |
| **Standard deviation** | 21.6 | 27.8 |
| **Range** | 0.6 to 79.3 | 18.2 to 63.5 |

**Supplementary Table 2.** Phylum, species and strains of bacteria used as antigenic stimuli

| **Species** | **Strain** | **Phylum** |
| --- | --- | --- |
| *Schaalia turicensis* | L12-BSM4 | *Actinomycetota* |
| *Bacillus licheniformis* | 4.60 | *Bacillota* |
| *Bacteroides ovatus* | DSM 1896^T^ | *Bacteroidota* |
| *Bifidobacterium longum* | L10-BSM5 | *Actinomycetota* |
| *Bifidobacterium pseudocatenulatum* | L22-MRS2 | *Actinomycetota* |
| *Citrobacter gillenii* | L26-FAA1 | *Pseudomonadota* |
| *Clostridium paraputrificum* | L16-FAA6 | *Bacillota* |
| *Collinsella aerofaciens* | D1-33 | *Actinomycetota* |
| *Enterococcus gallinarum* | L18-MRS5 | *Bacillota* |
| *Escherichia coli* | L1-FAA1 | *Pseudomonadota* |
| *Hafnia paralvei* | L15-FAA9 | *Pseudomonadota* |
| *Klebsiella pneumoniae* | L4-MRS2 | *Pseudomonadota* |
| *Lactiplantibacillus plantarum* | L16-MRS1 | *Bacillota* |
| *Lacticaseibacillus rhamnosus* | L24-FAA6 | *Bacillota* |
| *Parabacteroides merdae* | L18-MRS4 | *Bacteroidota* |
| *Staphylococcus epidermidis* | L20-MRS10 | *Bacillota* |
| *Streptococcus parasanguinis* | L12-MRS7 | *Bacillota* |
| *Sutterella wadsworthensis* | L15-FAA3 | *Bacillota* |
| *Veillonella atypica* | L12-MRS2 | *Bacillota* |

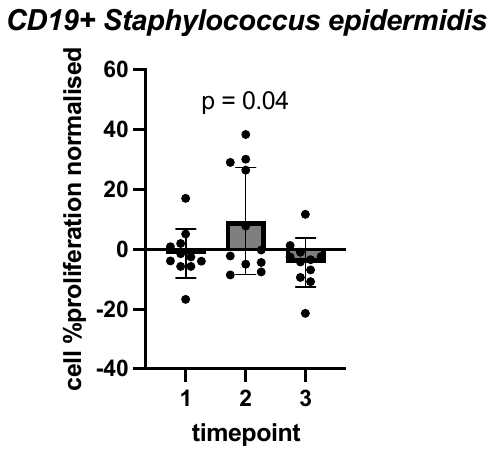

**Supplementary Figure 1.** Impact of DFg on circulating CD19^+^ cells. PBMCs (*Staphylococcus epidermidis*, CD19^+^) from patients with CRC and ileostomy undergoing DF regimen over 8 weeks (time points: 1, pre feeding; 2, 3 weeks; 3, 8 weeks), n=12. Cells exposed to antigenic stimulation and cultured *in vitro*. Data normalised via comparison with control well and analysed using one way ANOVA.

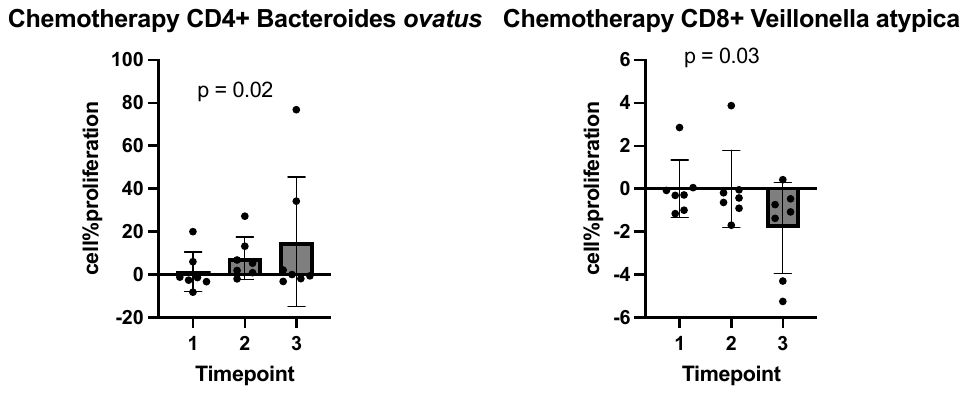

Chemotherapy CD4^+^ *Bacteroides ovatus*	Chemotherapy CD8^+^ *Veillonella atypica*

**Supplementary Figure 2.** Impact of chemotherapy on PBMC proliferation when exposed to bacterial stimuli in patients with CRC and ileostomy undergoing DF. PBMCs (CD4^+^/CD3^+^, CD8^+^) from patients with CRC, ileostomy and chemotherapy undergoing DF regimen over 8 weeks (time points: 1, pre feeding; 2, 3 weeks; 3, 8 weeks), n=6. Cells exposed to antigenic stimulation and cultured *in vitro*. Data normalised via comparison with control well and analysed using one way ANOVA.

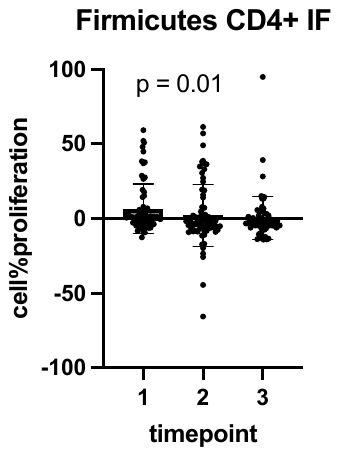

**Supplementary Figure 3.** Taxon-based analysis of the impact of DF on PBMC proliferation (*Bacillota*, CD4^+^) *in vitro*. PBMCs from patients with IF and stoma undergoing DF regimen over 8 weeks (time points: 1, pre feeding; 2, 3 weeks; 3, 8 weeks). Cells exposed to bacterial antigenic stimulation and cultured *in vitro*. Proliferative responses to bacteria measured, normalised compared to a control well and analysed using one way ANOVA.

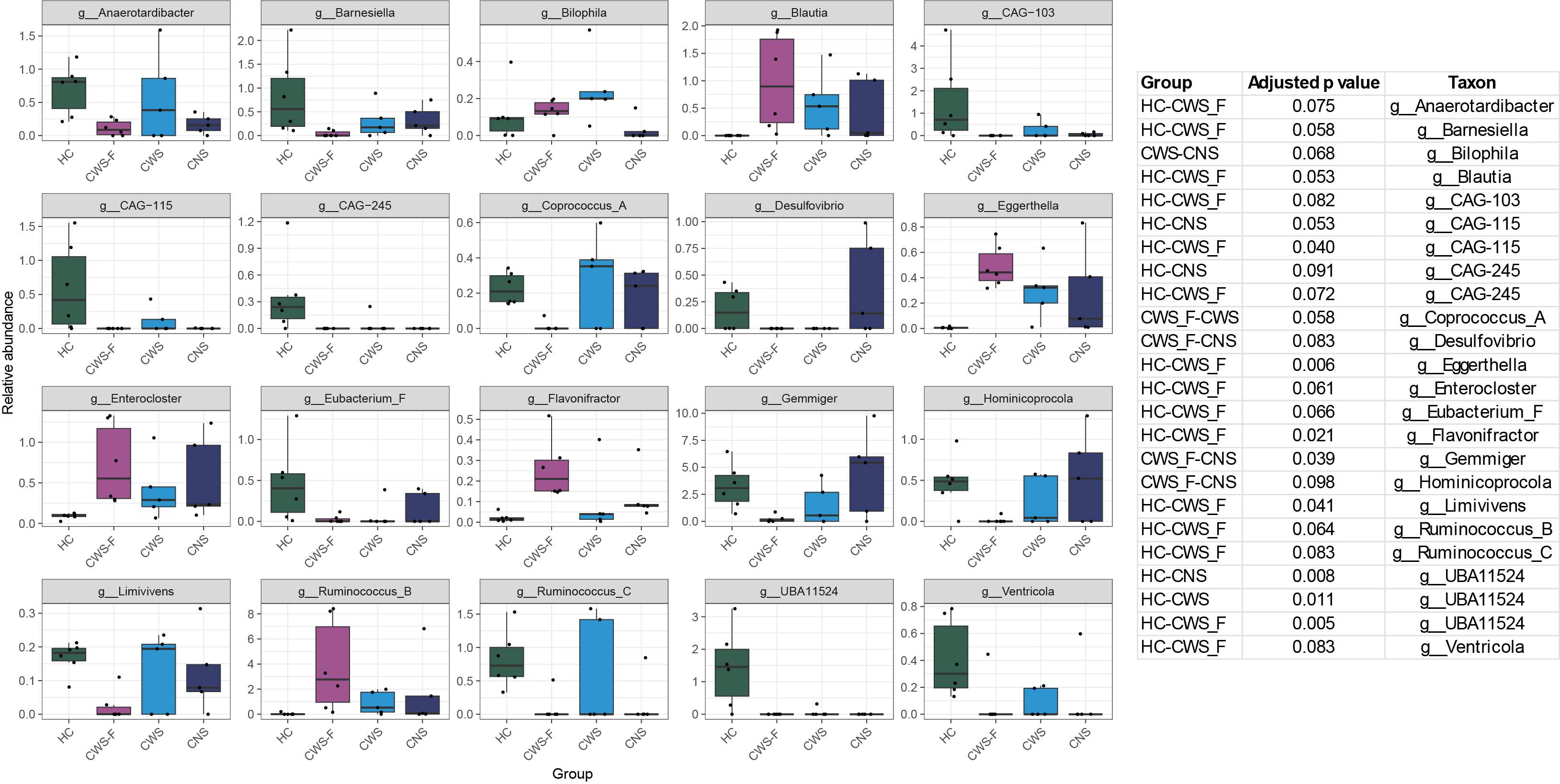

**Supplementary Figure 4.** Summary of genus-level data showing significant (FDR Padj < 0.1) differences between one or more patient groups. HC, healthy control (n=6); CWS-F, cancer with stoma and DF (n=6); CWS, cancer with stoma (no DF) (n=5); CNS, cancer no stoma (n=5). One-way ANOVA with Tukey’s post hoc testing was used to compare groups to one another (significant adjusted p values shown in the table to the right of the images).

**METHODS**

**Cohort description**

Fifty healthy, colorectal cancer (CRC) and intestinal failure (IF) patients were recruited from multiple hospital sites within London North West University Healthcare NHS Trust. An additional group of IF patients was compared retrospectively (n=55) to compare clinical outcomes as restoration of continuity surgery in this group of patients is more complicated than standard reversal of stoma (ROS), so could not be standardized.

Patients with anastomotic leak, inaccessible ileostomies and bowel strictures were excluded via water soluble enema and, if required, fistulogram. The CRC group of patients had undergone a form of rectal cancer resection called total mesorectal excision (TME) at anatomically similar locations and were defunctioned at the same level via ileostomy. They were all fed with the same distal feeding (DF) diet: Vital 1.5™, a nutritional drink (Abbott nutrition, 1.5 kcal/ml complete, balanced, peptide-based liquid) was used for DF in all recruited CRC patients, for an 8-week regimen, concluding with ROS, this was discussed and elucidated in patient participation exercises prior to commencement of the study.

In the IF group two feeding protocols were used: re-infusion of small bowel effluent and infusion of Vital 1.5, the latter being the standard DF regimen of choice used by St Mark’s Hospital. Chyme feeding was limited to patients following IF surgery alone, as focus groups for patients following CRC surgery suggested that most patients would not tolerate it and found it unacceptable conceptually.

The IF patients had heterogenous operations with either stomas or enterocutaneous fistulas at different locations of bowel, and one third of this cohort had inflammatory bowel disease. One third of these patients were fed with their own small bowel effluent (chyme), the remainder had the same diet regimen as the cancer cohort. Patients were selected for chyme by the IF team at St Mark’s Hospital.

We matched a CRC DF cohort (n=12 age range 41–86 years), an IF DF cohort (n=14, age range 22–79 years) with comparable control groups including CRC patients who were not distally fed (n=11) and healthy controls (n=9), CRC patients prior to their initial surgical resection (n=8), non-DF IF controls (n = 4) and with healthy controls (n=9). Control patients donated blood and stool only. None of the healthy control patients had any other pathology, were not taking any medication at the time and were fit and well at time of blood donation.

Chemotherapy/radiotherapy subgroup analyses were necessary to determine the separate impact on microbiota, serum metabolome and immune responses. All patients filled in dietary questionnaires prior to recruitment and any sample acquisition.

All recruited patients were offered a course of DF for 8 weeks, and all CRC patients but one underwent ROS thereafter. They were subsequently followed-up 6 months post-operatively. The IF patients were more variable as the extent of their continuity surgery was more individualized, therefore prospective clinical and mechanistic assessments for this cohort were only taken during the DF regimen and not post operatively.

**Ethics**

Collection and analysis of human samples, consent for qualitative interviews and collection of clinical data received approval from Research and Development at London North West University Trust and from the UK National Research Ethic Centres (IRAS 248290) in May 2018. Written informed consent was received by participants prior to inclusion in the study.

**Recruited distally fed patients**

Recruited patients for DF gave blood and urine samples pre-DF, alongside stoma effluent and bowel biopsies, taken 10 cm downstream from their stoma/fistula. All patients filled out qualitative questionnaires pre-DF, and a selection gave pre-DF qualitative interviews.

Patients subsequently gave blood samples and qualitative questionnaires at week 3 of DF and, at week 8 of the DF regimen donated blood and bowel biopsies from 10 cm distal to their stoma/fistula and repeated their qualitative questionnaires. Six months following closure of ileostomy, the cancer patients repeated their blood tests and donated stool samples which were stored at -80 °C until processed for shotgun metagenomics.

**Blood collection for immunology**

Whole blood was collected from patients in 3 heparinized 10 ml sterile glass vials and contemporaneously analysed.

**Microbial samples**

**Faeces**

Faecal samples were collected from patients in sterile 15 ml cryovials (of which one was stored (empty) as a negative control) and then snap frozen in liquid nitrogen immediately and stored at -80 °C. These samples were randomized prior to extraction of DNA. Faecal samples were defrosted and DNA was extracted using a DNeasy PowerSoil Pro Kit (Qiagen). DNA was quantified using Qubit with a broad-range DNA kit (ThermoFisher), then sent to Novogene (Cambridge, UK) for metagenomic (total DNA) sequencing.

**Tissue**

Limited enteroscopies to acquire samples were performed by expert endoscopists at St Mark’s Hospital. Six endoscopic biopsies were taken under direct vision, 10 cm from the stomal orifice to avoid predominately adjacent facultative aerobic species. Two were stored in formalin (10 %) for histopathology. Samples were then stored at room temperature, randomized and analysed in three batches to ensure that there were no biases between timepoint, fixation and section. Blinded tissue analysis was undertaken by a senior histopathologist and reported appropriately with diversion colitis graded to allow for analysis. Individual cases were discussed subsequently to take pictures and to discuss the implications of grading. These data were subsequently compared with clinical data taken at these time points regarding signs and symptoms of diversion colitis.

**Distal limb microbiota**

Distal limb microbiota samples were brushed from 10 cm distal to the stomal opening in the distal limb using a standard cytology brush and are collected directly into a sterile 2 ml cryotube. These were snap-frozen immediately and then stored in a -80 ° C freezer until processed for metataxonomic analysis.

**Serum samples for metabolomics**

A blood sample (10 ml) was taken in a standard red top bottle with a clot activator and allowed to clot for 15 min. Following this, the sample was centrifuged for 15 min at 1600 ***g***, at 4 °C. The resulting supernatant was aliquoted into 1.8 ml Eppendorf tubes, 400 µl per tube and frozen at -80 °C until analysis.

**Immunology**

Peripheral blood mononuclear cells (PBMCs) were collected from patients and inoculated with microbial antigenic material to examine memory T and B cell responses and compared with both healthy controls and non-healthy controls. T and B cell proliferative responses were assessed by flow cytometry using cell trace violet staining and antibodies to CD3^+^, CD4^+^, CD8^+^ and CD19^+^ cells to identify T and B cells, along with antibodies to gut and skin homing markers to demonstrate tissue localization using our previously published protocols.

Fresh PMBCs were obtained from 30 ml blood. Plasma was also stored at -80 °C for further analysis. PBMCs were labelled with CellTrace violet dye (CTV) at a concentration of 5 µM in PBS according to kit instructions (Life Technologies). CTV-labelled PBMCs were resuspended at 4x10^6^/ml in XVIVO15 medium supplemented with 1 % L-glutamine, 0.1 % gentamicin and 1 % penicillin/streptomycin. Labelled PBMCs were plated in a sterile flat-bottomed 96-well plate at 8x10^5^ cells/well in 200 µl of medium and inoculated with 2x10^5^ heat-killed bacteria (see **Supplementary Table 2** for strains).

**Bacterial species**

Nineteen species of bacteria isolated from the gastrointestinal tract were included in the study and used as described previously (Noble *et al.*, 2020). Two positive controls (staphylococcal enterotoxin B - SEB, a superantigen for CD4/CD8 T cells) and CpG oligodeoxynucleotide (CpG, a strong B cell inducer) and two negative controls (untreated cells and cyclosporin A) were used to validate a positive proliferative response. Following 7 days’ incubation with bacteria at 37 °C, 5% CO_2_, the patients’ PBMC proliferation was analysed by flow cytometry as described previously. Cell viability was determined using NearIR fixable live/dead stain (Life Technologies, 1:1000), according to manufacturer’s instructions. Cells were then incubated with fluorochrome conjugated antibodies.

Subsequently, to confirm that all CD4^+^ cells were T cells a CD3 conjugated antibody was included for 10 patients, replacing integrin ß7. Cells were fixed in 1 % paraformaldehyde solution (PFA) and stored at 4 °C until acquisition of data on a FACS Canto II (BD).

Data analysis was performed using FlowJo software (Version 10). Lymphocytes were identified using FSC/SSC properties followed by gating on single live cells and then B (CD19^+^) and T (CD4^+^ or CD8^+^) cells. Finally, dilution of CTV staining was used to determine the amount of proliferation. If cells had proliferated more than twice the amount of the negative control, it was considered a positive response.

**Analysis**

Results were analysed using GraphPad Prism 9 software (GraphPad, San Diego, CA) with comparisons made between time points of feeding per bacterial species, disease aetiology (CD, IF due to trauma or mesenteric ischaemia or CRC), treatment modality (chemotherapy and non-chemotherapy), between bacterial taxa and species.

Analyses were made of divided cells in CD4^+^, CD8^+^ and CD19^+^ cells, respectively, with additional analyses made of undivided cells when considering gut and skin homing to assess cells expressed when cells proliferated in response to bacterial antigen. Background cell proliferation was removed using the below equation.

***a = p – 2n*** *(a = adjusted value, p = cell proliferation, n = negative control)*

Adjusted cell proliferative data were tested for normality using Q-Q plots and analysed using one-way and two-way analysis of variance (ANOVA) and two-tailed paired *t* tests. P values <0.05 were considered significant unless multiple comparisons were made, in which case a new Bonferroni calculation was used.

If results were non-parametric, analyses were made using Friedman’s and Kruskall-Wallis tests.

**Specific analysis for pre and post cancer resectional data**

PMBC isolated from peripheral blood of eight patients pre-CRC resection (50 % chemotherapy/radiotherapy) were compared with post cancer resection patients (with formation of ileostomy) and with healthy controls after culturing with 11 species of common caecal bacteria for 7 days. This was a reduced panel of bacteria as patients prior to cancer resection have reduced cell counts.

Statistical comparisons were made between CD4+, CD8+ and CD19+ cells using ANOVA and post hoc ANOVA testing (Tukey tests), after determination of Gaussian distribution using Q-Q plots. These comparisons were used to determine whether there was a difference in means between pre resection, post resection and ileostomy and with healthy control groups, with the post hoc testing to determine exactly where the differences were between groups. All values were measured using the equation detailed above, which removes background PBMC proliferation from the negative control well in the experiment.

**Statistical comparisons in IF and cancer**

Comparisons were made between total proliferative responses between timepoints, bacterial phyla proliferative responses and direct comparisons between cell proliferative responses to individual species. Initial comparisons were made between healthy controls and pre fed values, to demonstrate the defunctioned proliferative response in benign patients. For these unpaired two tailed T tests for the normalised data were used. Corrected analysis of statistical significance using the Bonferroni method were used p value for significance 0.025 (using the equation 1(1-**α**)/T).

As the raw changes in DF proliferation are not normally distributed, non-parametric Friedman testing for these wide comparisons was used to compare ranks, and then post hoc testing (Dunn) to determine the changes between time-points. Unmatched samples (patient drop-outs) were removed to enable cross-timepoint analysis without formation of an artificial mean result. However excluded results were used when making single unmatched timepoint comparisons (e.g. timepoint 1 vs timepoint 3) to allow the use of these data.

For all comparisons beyond overall changes, normalised data were used, using each patients’ negative control well of cells. For phylum-specific results, the normalised data were parametric, therefore more conventional ANOVA and Tukey testing was used, and for species-specific comparisons. Finally, gut and skin homing responses were compared in this group in response to DF - to determine the systemic impact of DF on proliferating communities of PBMC *in vivo*.

**16S rRNA gene-based metataxonomic analysis**

Samples were prepared for DNA extraction using a modified phenol/chloroform extraction method (<https://doi.org/10.17504/protocols.io.bf28jqhw>). Sequencing was performed on an Illumina MiSeq platform (Illumina Inc., Saffron Walden, UK) using the MiSeq Reagent Kit v3 (Illumina) and paired-end 300 bp chemistry. The resulting sequencing data were processed using RStudio, version 1.3.1056 following the DADA2 v.1.18.0 pipeline as described previously (Callahan et al., 2016). Sequence alignments were performed using the Genome Taxonomy Database (GTDB) v86 ([https://gtdb.ecogenomic.org](about:blank)). Immediately after DADA2, the decontam package (Davis et al., 2018) was used to identify and remove possible contaminants present in the samples. As a result, an average of 28,277 reads per sample were obtained, ranging from 1,125 to 195,962 reads per sample.

Non-rarefied counts were subject to analyses (Wilcoxon rank sum test for comparisons of control) using ALDex2 v.1.18.0 (Fernandes *et al.*, 2014), microbiome (Lahti *et al*., 2017) and phyloseq (McMurdie & Holmes, 2013). Packages were used to calculate the different alpha diversity indexes for all samples. Statistical analysis was performed by 1-way ANOVA or Kruskal-Wallis tests. Statistically significant differences were then evaluated by Tukey or Wilcoxon rank-sum post hoc tests. Beta diversity, heatmaps and LEfSe analysis were obtained with the web-based statistical tool MicrobiomeAnalyst (Chong *et al.*, 2020).

**Shotgun metagenomic analysis of faecal microbiota**

Shotgun metagenomic sequence data were generated by Novogene (NovaSeq 6000; PE150 strategy). The sequence data returned to us were checked using fastQC v0.11.9 ([https://www.bioinformatics.babraham.ac.uk/projects/fastqc/](about:blank)), with an average of 67.5 Gb (+/- 5.8 Gb) generated for each sample. No trimming of data was required. Human DNA within samples was detected by mapping reads against the human genome (GRCh38/hg38; [https://hgdownload.soe.ucsc.edu/downloads.html](about:blank)) using bwa mem v0.7.17-r1188 (Li, 2013). Non-human DNA was extracted from read files using samtools v1.10 ([http://www.htslib.org/](about:blank)). Data were de-duplicated using FastUniq v1.1 (Xu et al., 2012). These human-filtered, de-duplicated files were uploaded to the European Nucleotide Archive and are available under BioProject PRJEB38258, and were used for all subsequent analyses.

Due to the large size of the human-filtered, deduplicated read files (total read pairs in each dataset representing 54.3 +/- 4.4 Gb), Megahit v1.2.9 (Li et al., 2016) was used to assemble sequence data for each of the 24 datasets. Unassembled reads were then pooled and subjected to a second round of assembly to improve the representation of low-abundance sequences (Hoyles et al., 2018). Genes in assemblies were predicted using MetaGeneMark (Besemer & Borodovsky, 1999; Zhu et al., 2010). Predicted genes were translated, and the protein sequences clustered using UCLUST (Edgar, 2010) with a 95 % cut-off identity (Hoyles et al., 2018).

Centroid sequences from each cluster were used to generate a non-redundant gene catalogue for determination of gene abundances and functional predictions. Gene abundances in each sample were determined as described previously (Hoyles et al., 2018). eggNOG-mapper v2 (eggNOG 5.0) was used to generate functional predictions for the dataset (Cantalapiedra et al., 2021): 2,400,538 genes had KEGG Orthology terms associated with them; 3,679,772 with COG terms; 691,700 with GO terms; 91,280 with CAZy terms. Taxonomic abundance data for archaea and bacteria were generated using sylph v0.8.1 (Shaw & Yu, 2025) and the pre-compiled GTBD_r220 sylph index.

Microbial gene richness was determined as described previously (Le Chatelier et al., 2013; Hoyles et al., 2018). Data were downsized to adjust for sequencing depth and technical variability by randomly selecting 20 million reads mapped to the merged gene catalogue (of 6,130,055 genes) for each sample and then computing the mean number of genes over 30 random drawings.

The groups were divided and statistically analysed after removal of an outlier (patient a10; **Figure A**): CNS, n=5; CWS, n=5; CWS-F, n=6; HC, n=7).

Metabat2 v2.12.1 (Kang et al., 2019) was used to bin assembled contigs. CheckM v1.0.18 (Parks et al., 2015) was used to determine contamination and completeness of the bins. Of a total of 6,889 across the 24 patient samples and the pooled sample, 1,274 bins with ≥80 % completeness and ≤10 % contamination were processed using MAGpurify v2.1.2 (Nayfach et al., 2019), then they were dereplicated using dRep v3.2.0 (Olm et al., 2017) with CheckM v1.0.18. This left 960 dereplicated metagenome-assembled genomes (MAGs) (available from [figshare](https://figshare.com/projects/Feeding_study_-_Dilke_et_al_/121884); **Figure B**). Identities of MAGs were determined using sourmash (Pierce et al., 2019) with GTDB r220 (Parks et al., 2022). A phylogenetic tree incorporating the 960 dereplicated MAGs was generated using PhyloPhlAn 0.99 (Segata et al., 2013) and visualized using iTOL v7 (Letunic & Bork, 2024). Overall quality of MAGs was assessed according to Bowers et al. (2017): those encoding 23S, 16S and 5S rRNA genes and at least 18 tRNAs (Prokka-annotated, v1.13.0; Seemann, 2014) with ≥90 % completeness and ≤5 % contamination were considered high-quality draft genomes; those with ≥50 % completeness and ≤10% contamination were considered medium-quality draft genomes; low-quality draft genomes are not reported on.

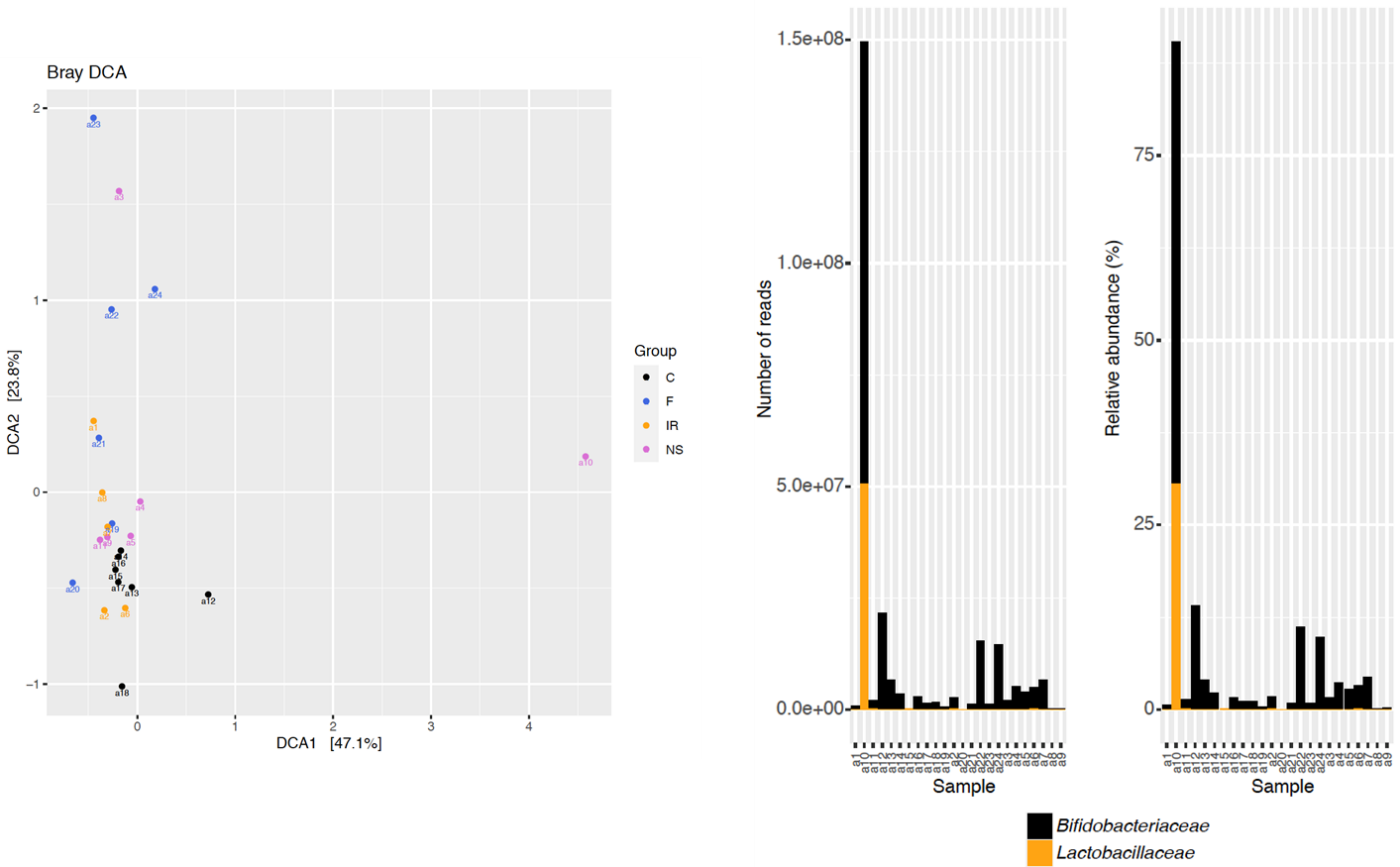

**Figure A.** Patient a10 was identified as an outlier in initial shotgun metagenomic data. Their faecal microbiota was almost entirely dominated by bacteria belonging to the families *Lactobacillaceae* and *Bifidobacteriaceae* in comparison with the other samples; hence, this sample was excluded from comparative analyses (it was included in the generation of MAGs).

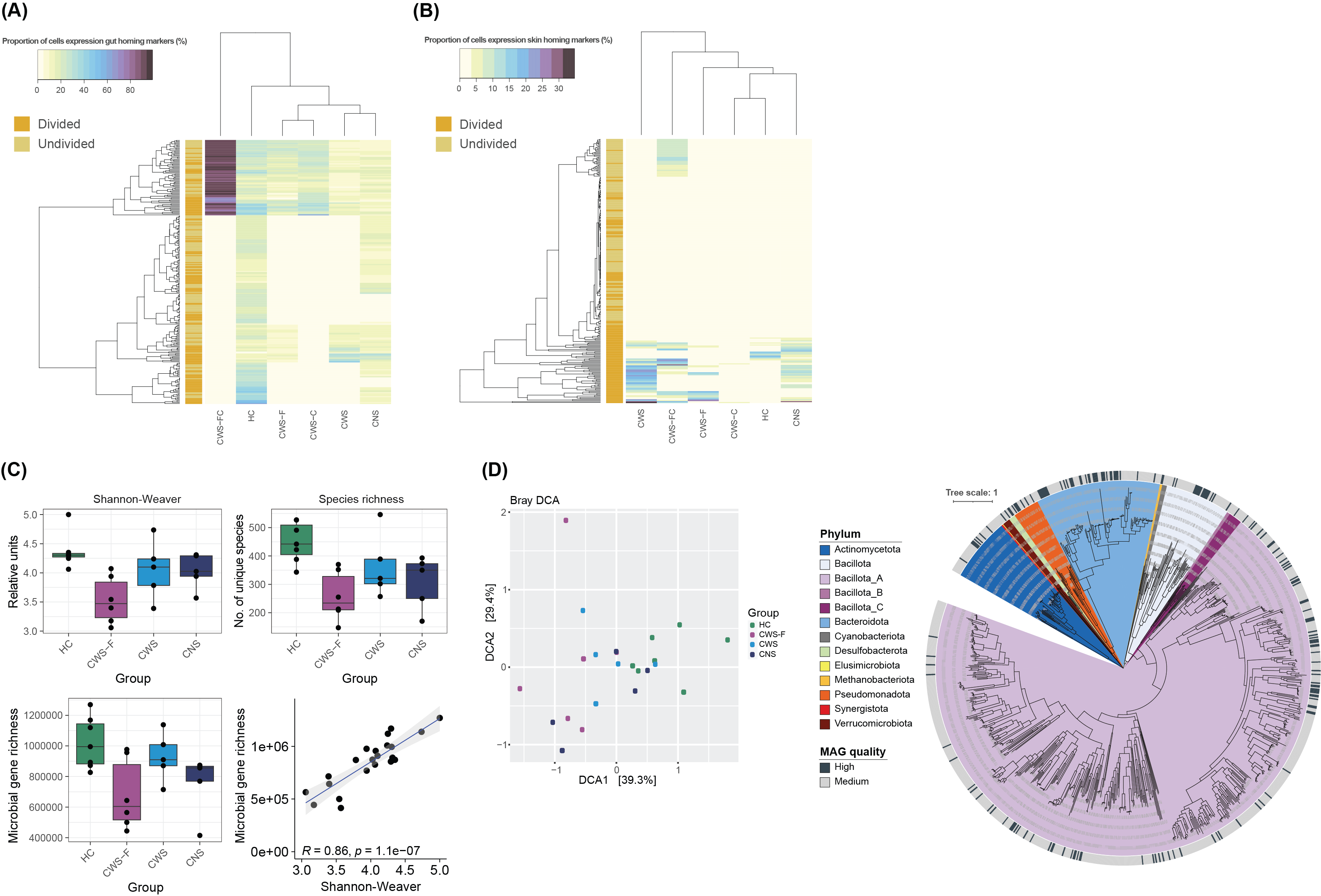

**Figure B.** Metagenome-assembled genomes were generated from the shotgun metagenomic data of all patients included in this study.

**Mass spectrometry**

**LC-MS amino acid and biogenic amine determination, FIA-MS determination of carnitines, glycerophospholipids and sphingolipids**

All metabolites were determined by liquid chromatography tandem-mass spectrometry (LC-MS/MS) or flow injection analysis-tandem mass spectrometry (FIA-MS/MS), using a SCIEX 4000 QTRAP mass spectrometer (SCIEX, Toronto, Canada). These determinations were performed using AbsoluteIDQ® p180 (Biocrates Life Sciences AG, Innsbruck, Austria), a targeted metabolomics tool that includes internal standards and standard curves for up to 180 metabolites including hexoses, amino acids, biogenic amines, acyl-carnitines, glycerophospholipids and sphingolipids.

Concentrations for metabolites were determined using the MetIDQ™ software package, which is an integral part of the AbsoluteIDQ® kit. Briefly, standards, internal standards, quality controls and 10 μl of each serum sample were loaded on a 96-well filter plate for protein precipitation, derivatized on the filter with phenyl isothiocyanate (Sigma Aldrich, Milwaukee, WI, USA) and dried under nitrogen flow. Metabolites were extracted by centrifugation to a lower 96-well collection plate with extraction solvent (Biocrates Life Sciences AG, Innsbruck, Austria) and different aliquots of the extract were used for LC-MS/MS and FIA-MS/MS.

**LC-MS bile acid determination**

Bile acid determination was performed using an in-house reverse-phase MRM method (Imperial College London), including calibration curves for 21 bile acids, and 5 bile acid ^13^C-labelled internal standards. Aliquots (50 μl) of standards, blanks and of each serum sample were transferred to microcentrifuge tubes and internal standard mix was added to all of them. After addition of ice-cold methanol and centrifugation, supernatant was collected and transferred into glass vials, dried under nitrogen, resuspended in water and measured by LC-MS.

**Description of metabolite classes**

The measured serum metabolites and metabolite ratios were grouped into different classes:

1)	Amino acids and biogenic amines

2)	Bile acids

3)	Acyl-carnitines

4)	Lysophosphatidylcholine (LysoPC)

5)	Phosphatidylcholine alkyl-acyl (PC aa)

6)	Phosphatidylcholine alkyl-ether (PC ae)

7)	Sphingomyelins (SM)

8)	Metabolite groups and ratios

The statistical analyses were performed separately for each class. Multiple testing corrections were applied within each class (FDR). Multiple testing corrections were not applied to metabolite groups and ratios. Median normalisation, generalised log transformation, and auto scaling (mean-centered and divided by the standard deviation of each variable) were applied to data from groups 1-7, unless otherwise stated. No normalisation was applied to metabolite groups and ratios.

Le Chatelier E, Nielsen T, Qin J, Prifti E, Hildebrand F, Falony G, Almeida M, Arumugam M, Batto JM, Kennedy S, Leonard P, Li J, Burgdorf K, Grarup N, Jørgensen T, Brandslund I, Nielsen HB, Juncker AS, Bertalan M, Levenez F, Pons N, Rasmussen S, Sunagawa S, Tap J, Tims S, Zoetendal EG, Brunak S, Clément K, Doré J, Kleerebezem M, Kristiansen K, Renault P, Sicheritz-Ponten T, de Vos WM, Zucker JD, Raes J, Hansen T; MetaHIT consortium; Bork P, Wang J, Ehrlich SD, Pedersen O. Richness of human gut microbiome correlates with metabolic markers. Nature. 2013 Aug 29;500(7464):541-6. doi: 10.1038/nature12506. PMID: 23985870.

Letunic I, Bork P. Interactive Tree of Life (iTOL) v6: recent updates to the phylogenetic tree display and annotation tool. Nucleic Acids Res. 2024 Jul 5;52(W1):W78-W82. doi: 10.1093/nar/gkae268. PMID: 38613393; PMCID: PMC11223838.

Li D, Luo R, Liu CM, Leung CM, Ting HF, Sadakane K, Yamashita H, Lam TW. MEGAHIT v1.0: A fast and scalable metagenome assembler driven by advanced methodologies and community practices. Methods. 2016 Jun 1;102:3-11. doi: 10.1016/j.ymeth.2016.02.020. Epub 2016 Mar 21. PMID: 27012178.

Li H. Aligning sequence reads, clone sequences and assembly contigs with BWA-MEM. arXiv. 2013. <https://arxiv.org/abs/1303.3997>.

McMurdie PJ, Holmes S. phyloseq: an R package for reproducible interactive analysis and graphics of microbiome census data. PLoS One. 2013 Apr 22;8(4):e61217. doi: 10.1371/journal.pone.0061217. PMID: 23630581; PMCID: PMC3632530.

Nayfach S, Shi ZJ, Seshadri R, Pollard KS, Kyrpides NC. New insights from uncultivated genomes of the global human gut microbiome. Nature. 2019 Apr;568(7753):505-510. doi: 10.1038/s41586-019-1058-x. Epub 2019 Mar 13. PMID: 30867587; PMCID: PMC6784871.

Noble A, Durant L, Hoyles L, Mccartney AL, Man R, Segal J, Costello SP, Hendy P, Reddi D, Bouri S, Lim DNF, Pring T, O'Connor MJ, Datt P, Wilson A, Arebi N, Akbar A, Hart AL, Carding SR, Knight SC. Deficient Resident Memory T Cell and CD8 T Cell Response to Commensals in Inflammatory Bowel Disease. J Crohns Colitis. 2020 May 21;14(4):525-537. doi: 10.1093/ecco-jcc/jjz175. PMID: 31665283; PMCID: PMC7242004.

Parks DH, Imelfort M, Skennerton CT, Hugenholtz P, Tyson GW. CheckM: assessing the quality of microbial genomes recovered from isolates, single cells, and metagenomes. Genome Res. 2015 Jul;25(7):1043-55. doi: 10.1101/gr.186072.114. Epub 2015 May 14. PMID: 25977477; PMCID: PMC4484387.

Olm MR, Brown CT, Brooks B, Banfield JF. dRep: a tool for fast and accurate genomic comparisons that enables improved genome recovery from metagenomes through de-replication. ISME J. 2017 Dec;11(12):2864-2868. doi: 10.1038/ismej.2017.126. Epub 2017 Jul 25. PMID: 28742071; PMCID: PMC5702732.

Parks DH, Chuvochina M, Rinke C, Mussig AJ, Chaumeil PA, Hugenholtz P. GTDB: an ongoing census of bacterial and archaeal diversity through a phylogenetically consistent, rank normalized and complete genome-based taxonomy. Nucleic Acids Res. 2022 Jan 7;50(D1):D785-D794. doi: 10.1093/nar/gkab776. PMID: 34520557; PMCID: PMC8728215.

Pierce NT, Irber L, Reiter T, Brooks P, Brown CT. Large-scale sequence comparisons with *sourmash*. F1000Res. 2019 Jul 4;8:1006. doi: 10.12688/f1000research.19675.1. PMID: 31508216; PMCID: PMC6720031.

Seemann T. Prokka: rapid prokaryotic genome annotation. Bioinformatics. 2014 Jul 15;30(14):2068-9. doi: 10.1093/bioinformatics/btu153. Epub 2014 Mar 18. PMID: 24642063.

Segata N, Börnigen D, Morgan XC, Huttenhower C. PhyloPhlAn is a new method for improved phylogenetic and taxonomic placement of microbes. Nat Commun. 2013;4:2304. doi: 10.1038/ncomms3304. PMID: 23942190; PMCID: PMC3760377.

Shaw J, Yu YW. Rapid species-level metagenome profiling and containment estimation with sylph. Nat Biotechnol. 2025 Aug;43(8):1348-1359. doi: 10.1038/s41587-024-02412-y. Epub 2024 Oct 8. PMID: 39379646; PMCID: PMC12339375.

Xu H, Luo X, Qian J, Pang X, Song J, Qian G, Chen J, Chen S. FastUniq: a fast de novo duplicates removal tool for paired short reads. PLoS One. 2012;7(12):e52249. doi: 10.1371/journal.pone.0052249. Epub 2012 Dec 20. PMID: 23284954; PMCID: PMC3527383.

Zhu W, Lomsadze A, Borodovsky M. Ab initio gene identification in metagenomic sequences. Nucleic Acids Res. 2010 Jul;38(12):e132. doi: 10.1093/nar/gkq275. Epub 2010 Apr 19. PMID: 20403810; PMCID: PMC2896542.
